# Feasibility of real-time fMRI neurofeedback for rehabilitation of reading deficits in aphasia

**DOI:** 10.1101/2025.01.03.25319980

**Authors:** Olga Boukrina, Abubakar Yamin, Guang H Yue, Yekyung Kong, Yury Koush

**Author notes:** **Correspondence:** Olga Boukrina,.

## Abstract

**Background:** Reading impairments, a common consequence of stroke-induced aphasia, significantly hinder life participation, affecting both functional and leisure activities. Traditional post-stroke rehabilitation strategies often show limited generalization beyond trained materials, underscoring the need for novel interventions targeting the underlying neural mechanisms.

**Method:** This study investigates the feasibility and potential effectiveness of real-time functional magnetic resonance imaging (fMRI) neurofeedback (NFB) intervention for reading deficits associated with stroke and aphasia. We enrolled left-hemisphere stroke survivors in the subacute recovery period and healthy controls in an fMRI NFB intervention study focusing on increasing activation within the left supramarginal gyrus (SMG), a critical region for reading supporting orthography-phonology conversion.

**Results:** Preliminary findings demonstrate that stroke participants showed significant improvements in reading comprehension and phonological awareness, as evidenced by marked gains on the Reading Comprehension Battery for Aphasia (RCBA) and a phonology two-alternative forced choice test. Functional MRI results indicated that stroke participants exhibited increased activation from day 1 to day 3 of NFB training within the left SMG and the broader left hemisphere reading network, particularly during challenging nonword reading tasks. Healthy controls also showed increased activation during NFB regulation and reading tasks, but these changes were outside the traditional reading network, involving regions associated with cognitive control, reward anticipation, and learning. In both stroke participants and healthy controls, we also found changes in dynamic functional connectivity of multiple resting state networks from before to after NFB training.

**Conclusions:** Although preliminary, this research contributes to the development of biologically informed interventions for reading deficits in aphasia, representing an early step towards improving post-stroke rehabilitation outcomes. Future randomized controlled trials are necessary to validate these findings by including a sham NFB control group within a larger participant sample.

**Registration:** The study was preregistered on ClinicalTrials.gov, NCT# NCT04875936

## 1. Introduction

Among 795,000 new stroke survivors annually in the US and 13.7 million worldwide, 22% experience reading impairments as part of an acquired language disorder called aphasia^1^. For 61% of individuals with aphasia, language and reading deficits persist 1 year after stroke and negatively impact life quality^2^. In this study we explore the potential of real-time functional magnetic resonance imaging (fMRI) neurofeedback (NFB) to promote adaptive neural plasticity during subacute stroke and alleviate post-stroke language and reading impairments.

A major challenge in reading rehabilitation is the limited generalization of many treatments beyond trained materials^3^. Biologically inspired interventions, such as real-time fMRI NFB can overcome this limitation by directly promoting post-stroke neuroplasticity. By receiving feedback about their current brain state, participants can develop targeted mental strategies to regulate brain activity^4^. Repeated self-regulation efforts can drive learning-induced cognitive and motor improvements^5^.

Two studies have explored fMRI NFB for aphasia rehabilitation^6,7^. Sreedharan et al.^7^ trained 4 participants with subacute-to-chronic nonfluent aphasia (6 weeks to 6 months post-stroke) to regulate connectivity between Broca’s and Wernicke’s areas by performing covert language tasks over 4 sessions, resulting in increased connectivity between these regions. However, a follow-up study found no significant language improvements in the NFB group compared to usual care^6^. Thus, while real-time fMRI NFB shows potential in stroke rehabilitation by inducing neural changes, its impact on behavioral improvements needs to be further investigated.

The main goal of this study was to build on the relative success of previous fMRI NFB rehabilitation studies by developing a biologically inspired intervention for stroke-induced reading deficits. A critical part of achieving this goal is identifying specific brain mechanisms, which can be modulated to improve post-stroke reading ability.

A growing body of clinical neuroimaging literature in post-stroke aphasia suggests that in addition to the direct impact of stroke lesions on the left-hemisphere language and reading areas, there is persisting dysfunction in parts of the left hemisphere that are not directly affected by an obvious structural lesion. Multiple research teams have found reduced cerebral blood flow (CBF) and neural activity in perilesional areas and in functionally connected areas remote from the infarct zone^8–13^. Most of these studies also show that decreased CBF is negatively correlated with language performance. In our longitudinal study of 31 left-hemisphere stroke survivors^14^, both subacute (<5 weeks) and chronic (> 3 months post-stroke) regional CBF, measured with Arterial Spin Labeling (ASL), were reduced in the intact parts of the left reading network relative to their right-hemisphere counterparts. Our study lacked a non-stroke control group; however, a recent investigation of 43 chronic aphasia participants and 25 healthy controls confirmed a chronic reduction in left parietal and temporal CBF among individuals with aphasia that was correlated with residual language ability^12^.

Based on evidence linking left hemisphere hypoperfusion to persistent language deficits, we propose that timely restoration of cerebral perfusion and neural activity in intact left reading network components may enhance reading and language recovery. Consistently with this hypothesis, Hillis ^15^ showed that promoting reperfusion via a pharmacologically-induced increase in blood pressure in a patient with acutely hypoperfused left perisylvian language cortex transiently improved naming and comprehension. Saur *et al.*^16^ and Fridriksson^17,18^ reported that chronic aphasia recovery was associated with re-emergence of left brain activity. Finally, our perfusion study^14^, showed a positive linear relationship between subacute perfusion in the intact ipsilesional parts of the left reading network and chronic phonology competence, when controlling for lesion size.

While leveraging brain tissue directly damaged by stroke lesions is not feasible, fMRI NFB provides the opportunity to test whether self-regulating brain activity in functionally connected areas with reduced CBF can positively impact reading. To induce and maintain a left-dominant brain activation pattern, we combined fMRI NFB with right-hand motor imagery, a mental simulation of movement without motor output, which can be performed even by stroke patients with severe hand impairments. Lateralized motor imagery produces stronger activation in the hemisphere contralateral to the imagined movement (e.g., left hemisphere for right hand imagery), an asymmetry that can be further enhanced through fMRI NFB. We targeted the left supramarginal gyrus (SMG), a multimodal association area implicated in orthography-phonology conversion^19–22^, a skill important for reading and commonly impaired in left-hemisphere stroke patients^14,23,24^. In addition to its role in reading, left SMG is implicated in planning finger positioning for tool use^25^ functional grasps^26^, and kinesthetic motor imagery of finger movements^27^. Because of the SMGs dual role in motor planning and reading-related processes, we expected that right hand motor imagery could be an effective mental strategy to help increase its activation.

This paper focuses on the methodology and feasibility of fMRI NFB and motor imagery rehabilitation for reading deficits following left-hemisphere stroke. We evaluated various aspects of experimental design, including participant enrollment, retention, and motivation, the suitability of outcome measures, and the potential efficacy of the intervention.

## 2. Materials and Methods

### 2.1. Participants

Stroke participants were inpatients at an acute rehabilitation hospital screened using electronic review of medical charts. Neurotypical controls were recruited from the community. Four individuals with subacute left hemisphere stroke (*M age*=63.5, *SD*=17, all men, 2 Caucasian, 2 African American, *M days post stroke* =21.3, *SD* = 5.8, WAB 28 Aphasia Quotient (AQ)=73.38, *SD*=31.11) and 3 age-matched healthy controls (HCs) (*M age*=66, *SD*=6, all men, 2 Caucasian, 1 African American) agreed to take part in the study. All participants were right-handed, fluent and literate in English (prior to stroke), and had no contraindications to MRI. All stroke participants had a 1^st^ ever stroke (*M* lesion volume = 22.09 ml; *SD* = 23.57 ml) and post-stroke reading deficits, defined based on performance on subsets of the Reading Comprehension Battery for Aphasia–2^nd^ ed. (RCBA-2)^29^. According to the WAB, two stroke participants had anomic aphasia, one had transcortical motor aphasia, and one had Broca’s aphasia. Participants with a history of developmental dyslexia or neurological diagnoses, other than stroke and aphasia, were excluded. All participants with aphasia completed aphasia-friendly written informed consents. The study was approved by the Kessler Foundation Institutional Review Board. The feasibility data presented here are part of a larger ongoing study preregistered at ClinicalTrials.gov, NCT# NCT04875936.

### 2.2. Materials and Procedure

#### 2.2.1. Study Design

The study has a 2×2×3 mixed design, with participant group and fMRI NFB type (stroke vs HC; contingent vs. non-contingent) as between-subjects factors and session (T1, T2, T3) as a within-subjects factor. In the contingent fMRI NFB condition participants receive feedback based on their own brain activity, while in the non-contingent fMRI they receive yoked feedback based on brain activity of another participant. This control condition was selected to isolate the effect of feedback contingency, while keeping all other aspects of the study the same between conditions. In the feasibility data reported in this study all participants were assigned to the <u>contingent feedback</u> condition based on a pre-established randomization schedule. The first three stroke and HC participants were assigned to the contingent feedback condition, while subsequent participants were assigned to either condition based on a computer-generated random sequence. Participants were masked to their assignment, while outcome assessors were not. *A priori* power analysis indicated that given a medium effect size, 80% power for detecting interaction effects in each participant group (fMRI NFB type x treatment session) would be achieved with a sample of 28 participants (N=14 per fMRI NFB type). A medium effect size in this estimation represents a conservative estimate, as previous NFB studies with stroke patients reported medium to large effect sizes of fMRI NFB on brain activity^30,31^ (See supplemental materials for additional study details).

#### 2.2.2 Screening

Prior to randomization, participants underwent screening, including review of medical history, and completed demographic and handedness questionnaires^32^. Stroke participants additionally underwent screening for reading deficits using Paragraphs test (subtests VIII, IX) of the RCBA-229.

#### 2.2.3. Baseline and Outcome Assessments

Following randomization, participants underwent baseline testing, which included a comprehensive assessment of language impairments in aphasia using the WAB-Revised^28^, a reading comprehension assessment using the full RCBA-2, a picture naming test using the Boston Naming Test – short version^33^, and two-alternative forced choice (2AFC) touch screen computer tests of semantics, phonology, and orthography. Participants also completed a computerized reading aloud task where they were asked to read aloud 120 monosyllabic words and 80 length-matched pronounceable nonword letter strings. Nonwords were included because their novelty makes nonword reading more sensitive to phonological deficits, requiring effortful orthography-to-phonology mapping. Computer tests were presented on a 15-inch computer screen using OpenSesame software version 3.3.11^34^.

We assessed unilateral gross manual dexterity using the Box and Blocks test^35^, and grip strength using an electronic dynamometer (Jamar Smart Hand Dynamometer, Patterson Medical) to assess if motor imagery impacted motor function. All instruments were re-administered during outcome assessment.

The acceptability of the NFB intervention was assessed by self-report. These measures included motivation to participate in the study, commitment to the study, and perceived difficulty of the session, all rated on a 7-point Likert scale. For motivation and commitment, the scale was labeled as follows: 1 - very low, 2 - low, 3 - somewhat low, 4 - neutral, 5 - somewhat high, 6 - high, and 7 - very high. For difficulty, the scale was labeled as follows: 1 - very easy, 2 - easy, 3 - somewhat easy, 4 - neutral, 5 - somewhat hard, 6 - hard, and 7 - very hard.

#### 2.2.4. Neurofeedback Training

Participants were taught to engage in left-hand mental motor imagery exercises, including activities like finger tapping (random tapping of each right-hand finger to the thumb), typing (simulating keyboard use with each finger), playing piano keys, pointing, making gestures, or tracing text in a book with their finger. They were instructed to imagine a large amplitude of movement, adopting a first-person perspective. They were encouraged to imagine the accompanying sensory experience without moving their fingers. Participants completed the fMRI NFB task practice prior to beginning training.

After motor imagery and task practice, participants underwent MRI scans, including structural and functional scans (Fig.1). FMRI NFB training was completed over 3 sessions and included 2 15-minute runs per session. Each run had 9 fMRI NFB trials. We defined a trial as one whole cycle through the task sequence including feedback. Each trial lasted 96 seconds (Fig. 2). The trial began with a 21-second fixation screen to obtain baseline level of ROI activity followed by a 21-second mental motor imagery block. Participants then spent 24 seconds reading aloud words or pronounceable nonwords, after which they received a numeric feedback score accompanied by a smiley face for 4.5 seconds. The trial ended with a 22.5-second rest period, during which participants viewed a black screen. Task block presentation was locked to TR (1.5 seconds). Participants saw either 8 words or 8 nonwords during the reading task block, and this was randomized for each run and across training days. Each letter string was presented for 1 second followed by 1-3 second jitter. Voice responses were collected via an MRI compatible fiberoptic microphone (FOMRI III+, Optoacoustics) and used to score reading accuracy. For each NFB training session, the reading aloud stimuli consisted of 72 words and 72 nonwords. These included selections from both baseline and outcome assessments stimulus lists, as well as new items, matched across training days on linguistic properties.

**Figure 1.**
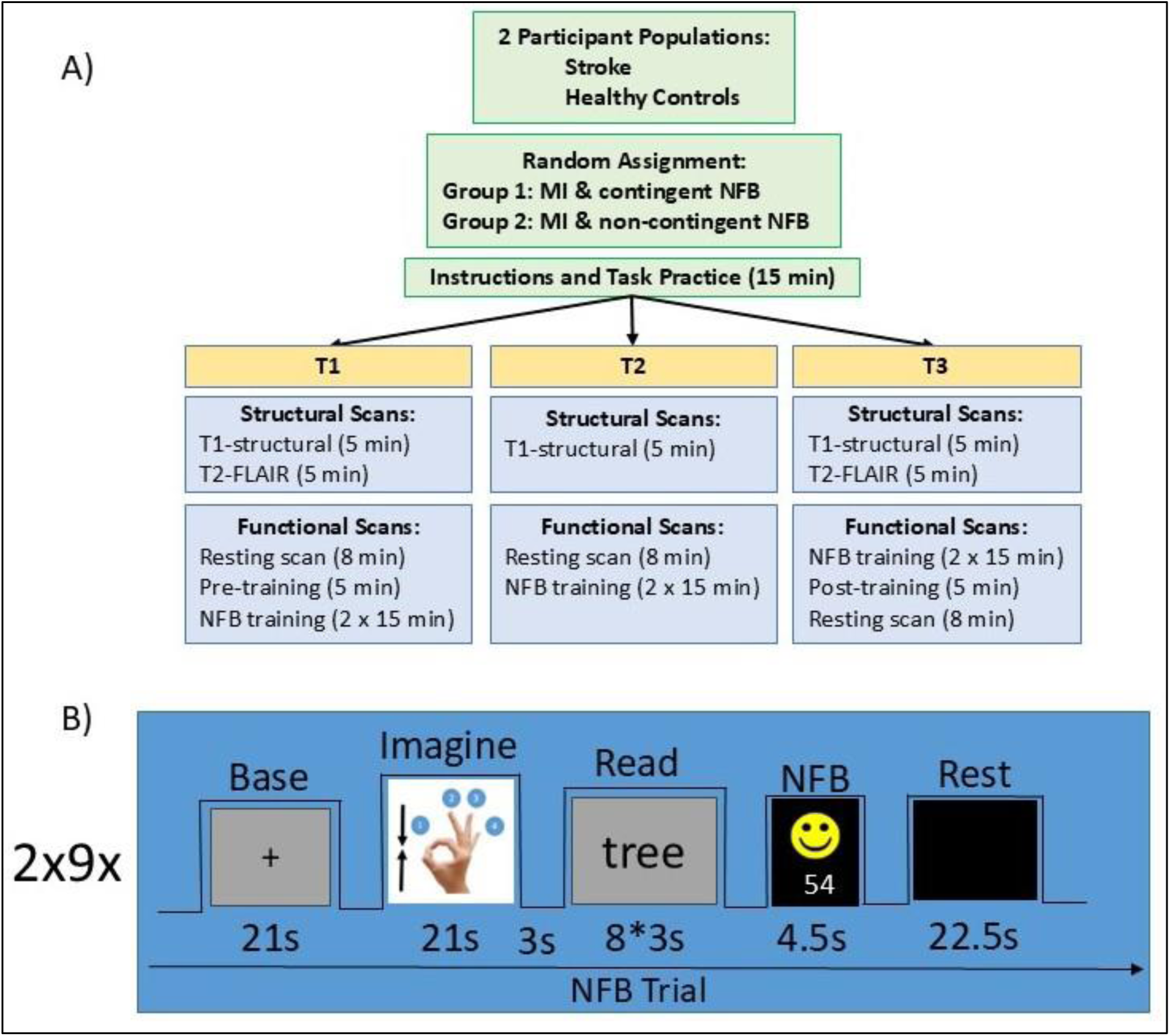
Study Design: A) Study timeline: Participants completed 3 weekly fMRI NFB training sessions labeled as T1, T2, and T3. At T1 and T3, we acquired Fluid Attenuated Inversion Recovery (FLAIR) scans to help delineate stroke lesions and monitor for new pathology. Prior to initiating and following completion of fMRI NFB training (T1 & T3) we acquired transfer runs (pre-training, post-training). These runs mirrored the training task without offering neurofeedback. T1-weighted structural images were acquired in every session to facilitate co-registration of atlas ROI to the participants’ brain. B): FMRI NFB Trial Structure. Each blocked trial started with a 21-second fixation period, followed by a 21-second regulation block, where participants were asked to engage in mental imagery. Next, participants read aloud words and pronounceable nonwords for 24 seconds and received numeric feedback (0-100) paired with a smiley face presented for 4.5s. The trial concluded with 22.5 seconds of rest indicated by black screen.

**Figure 2.**
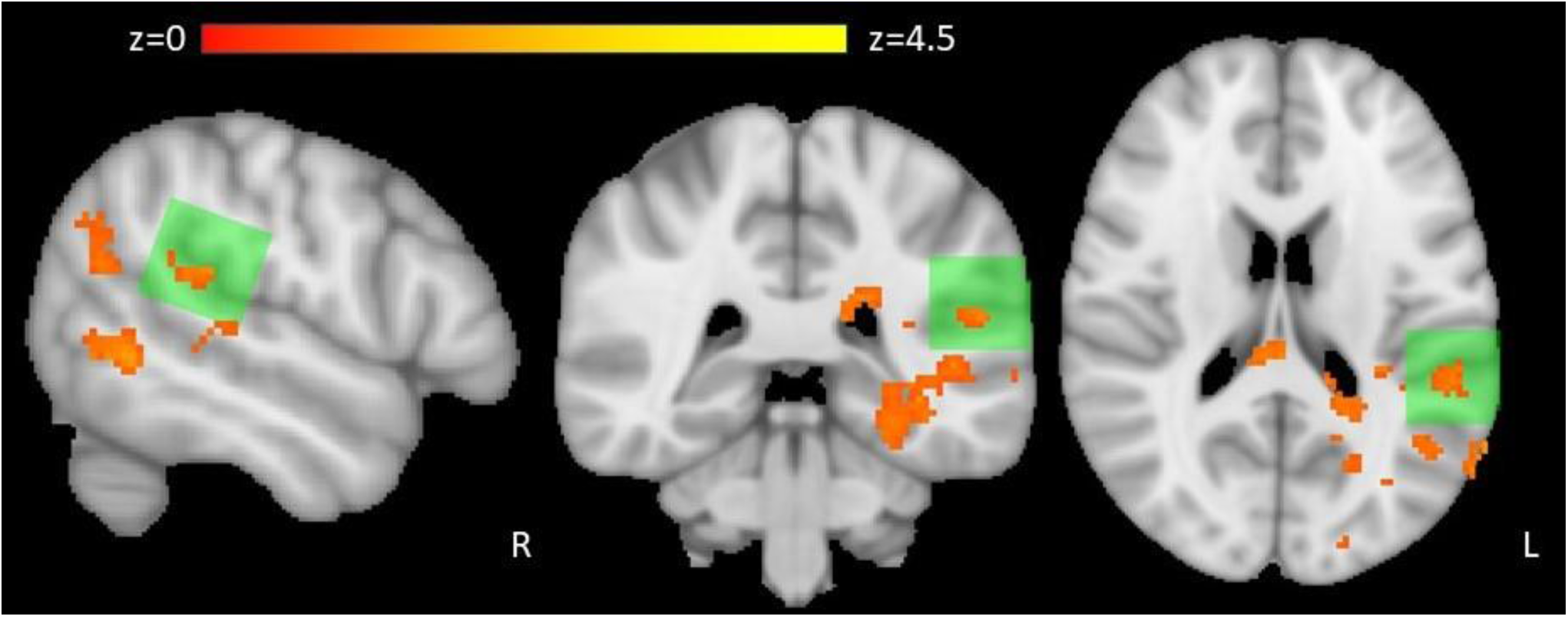
Brain activation increases among stroke participants during nonword reading relative to baseline on day 3 of NFB training compared to day 2. The target ROI used in NFB training is highlighted in green. Z (Gaussianised T/F) statistic images are displayed with a cluster threshold of z > 1.96, corrected to p = 0.05.

Online data analysis and stimuli presentation were implemented using OpenNFT (opennft.org). Participants received intermittent feedback based on percent signal change in the activity of the target ROI, scaled to 0-100 with negative values set to 0. NFB signal for each trial was computed as the difference between average activation in the “imagine” (MI) and “read” blocks and the average activation during baseline (+) blocks across the run, controlling for head motion. Higher positive feedback value paired with a cartoon smiling face corresponded to higher recruitment of the target ROI during motor imagery and reading blocks compared to baseline. All stimuli were back-projected onto an MRI-compatible screen and viewed by participants through a mirror attached to the head coil.

#### 2.2.5. Pre- and Post-Training Runs

NFB training was preceded and followed by pre- and post-training (transfer) runs (Fig.1). These runs were conducted according to the same procedure as each of the NFB training runs, except they consisted of 4 trials each. These scans were used to measure changes in activity from before to after NFB training and to confirm that NFB training could be transferred to a situation without feedback. The reading lists in the pre- and post-training runs included 32 additional words and nonwords (same stimuli for pre- and post-training runs) that were matched on all relevant linguistic properties to the stimuli presented during NFB training.

#### 2.2.6. Homework

Each participant completed 10 homework sessions, conducted outside of the scanner. Homework sessions mirrored the timing and nature of the NFB in-scanner sessions. They were presented using E-prime 3.0 software and were supervised by a research assistant. Identically to NFB runs (Fig. 2), each homework session consisted of 2 15-min runs with 9 trials each. While completing homework sessions, participants were encouraged to practice the imagery strategy that seemed to be most effective in generating high feedback scores during the in-scanner NFB training sessions. Homework sessions 1-5 were conducted on separate days after the 1st in scanner NFB training session. Homework sessions 6-10 occurred on separate days after the 2nd in scanner NFB training session.

#### 2.2.7. ROI mask selection

Left SMG was selected as the target ROI in this study for its key role in letter-to-sound conversion. To confirm that SMG is reliably activated by both reading and motor imagery, we conducted a pilot study in 6 healthy controls (HC, 4 women, 24-39 y.o.) (See Supplemental Materials).

#### 2.2.8. Neuroimaging

Participants underwent 3 MRI scans (Fig. 1, T1, T2, T3) on a 3.0T Skyra Siemens scanner using a 20-channel head coil. <u>Structural Neuroimaging</u>. To facilitate co-registration of atlas ROI to the participants’ brain we acquired T1-weighted structural scans (TR=2100 ms, TE=3.43 ms, TI=900ms, 176 sagittal slices, 1mm^3^ isotropic voxels) from each participant at each timepoint. In addition, to help segment stroke lesions and detect any new pathology, a T2-weighted Fluid Attenuated Inversion Recovery (FLAIR) scan (TR=9000 ms, TE=91 ms, TI=2500ms, 50 slices, 1×1×3mm^3^ voxels) was acquired at T1 and T3. <u>Functional Neuroimaging</u>. FMRI scans were acquired using rapid simultaneous multi-slice echo-planar imaging (EPI) (TR=1.5s, TE=30ms, 44 slices, gap = .5mm, 2mm^3^ isotropic voxels). Functional scans included resting state scans (326 TRs), pre-training (588 TRs), training and post-training scans (262 TRs). To standardize the rest condition across participants, we instructed participants to look at a centrally presented fixation dot for the duration of the scan and let their mind wander.

#### 2.2.9. Lesion Mapping

Lesions were labeled using a combination of manual segmentation and automated intensity-based voxel selection. To avoid warping of the lesion area during registration to the anatomical template, we applied cost-function masking of the input image using the inverse of the lesion mask (lesion weights). Lesion size served as a covariate in the analyses where appropriate.

#### 2.2.10. Analysis

##### Behavioral Data

The study was powered to detect group differences over time. Although this feasibility study did not include the non-contingent NFB group, we compared the effects of testing session (baseline vs. outcome) between healthy controls and stroke participants using a repeated measures analysis. Group was treated as a between-subjects factor and testing session as a within-subjects factor. We report p-values, effect sizes, and observed power for the group by testing session interaction. We also evaluated motivation and commitment to participation in the study, and difficulty of the current NFB session on each of the 3 MRI NFB training days, using a single sample t-test comparing each value to 1 (very low commitment and motivation/very easy). We expected that motivation and commitment would be significantly different from 1 while difficulty would not.

##### Task fMRI

FMRI data processing was carried out using FEAT (FMRI Expert Analysis Tool)^36^. The 8 fMRI runs per participant (2 pre- and post-training runs and 6 NFB training runs) were skull-stripped, motion corrected, smoothed at 6mm FWHM, and high pass filtered with a cutoff of 100s. Each run was co-registered with the participants’ T1-weighted anatomical images and with the standard space MNI152_T1_2mm_brain template. We modeled each event during the trial (baseline fixation, NFB regulation (motor imagery), word reading, nonword reading, and NFB display) as separate regressors. The model additionally included standard motion parameters and their temporal derivatives. For each run, we modeled the contrasts between the block of interest (NFB regulation, reading words, and reading nonwords) and the baseline. Whole brain group analyses were performed separately for stroke and healthy control participants and for NFB training and transfer runs. We anticipated activation increases in the target ROI and throughout the left hemisphere language network. For the NFB training runs, we contrasted consecutive runs to determine if brain activity was increasing as training progressed. For the transfer runs we compared post-training to pre-training runs. Group-level effects were estimated using FLAME (FMRIB’s Local Analysis of Mixed Effects) stages 1 and 2. Z (Gaussianised T/F) statistic images were thresholded using clusters determined by *z*>1.96 and a (corrected) cluster significance threshold of *p*=0.05^37^.

##### Resting State fMRI

Dynamic resting state functional connectivity (rsFC) was compared for rs-fMRI scans acquired before and after NFB training. Each rs-fMRI scan was skull stripped, motion corrected, smoothed with a 5-mm FWHM kernel, and aligned to the MNI152 2-mm brain template. Motion artifacts were further reduced using ICA-AROMA^38^. We additionally regressed out signals from CSF and WM, performed linear detrending and applied a Gaussian high-pass temporal filter, using Gaussian-weighted least-squares straight-line fitting with a 100-second FWHM. Lastly, we extracted mean timeseries from 333 regions of interest (ROI) in the Gordon *et al.* cortical area parcellation atlas^39^.

For each participant, we calculated dynamic functional connectivity (dFC) matrices using Pearson correlation. A sliding window of 60 TR (120 seconds) and a step size of 6 TR (12 seconds) was used, resulting in 44 dFC matrices per participant. Each dFC matrix was then recoded as a vector of geodesic distances to the centroids. These vectors were used to train a LASSO regularized logistic regression classifier with 5-fold cross-validation, aiming to identify variations in connectivity with the highest discriminatory power from before to after NFB training. Significance testing was performed using a permutation test, where the feature values were randomly shuffled between participants across groups, and only the features with weights exceeding the 95th percentile of the permutation test values were retained. We considered the top 1% of significant connections in the whole brain analysis.

## 3. Results

### Behavioral Data

There was a significant group-by-session interaction for the RCBA-2 and 2AFC phonology test. Relative to HCs, who were performing near perfect levels at both baseline and outcome testing sessions, stroke participants improved by 11 points on the RCBA-2, *F*(1,5)=8.5, *p*<0.05, *η^2^*=0.63, and by 15.42% on the 2AFC phonology test, *F*(1,5)=7.34, *p*<0.05, *η^2^*=0.59. No other behavioral tests showed significant differences (Table 1).

**Table 1.** Behavioral Baseline and Outcome Assessments, Including Means and Standard Deviations, P-values, Effect Sizes, and Observed Power.

| Test Name | Testing Session | Participant Group | | P-value (Group x Session Interaction) | Effect Size ($\eta^2$) | Observed Power |
| --- | --- | --- | --- | --- | --- | --- |
|  |  | HC | Stroke |  |  |  |
| RCBA (Max=100) | Baseline | 98.33 (2.08) | 72.5 (16.42) | 0.03* | 0.63 | 65% |
|  | Outcome | 98.33 (1.15) | 83.5 (18.08) |  |  |  |
| WAB (Max=100) | Baseline | 99.27 (0.42) | 73.38 (31.11) | 0.07 | 0.51 | 45% |
|  | Outcome | 98.80 (1.31) | 78.85 (27.87) |  |  |  |
| Reading Aloud Words (Max=100%) | Baseline | 98.67 (1.53) | 75.75 (39.2) | 0.36 | 0.17 | 13% |
|  | Outcome | 99.67 (0.58) | 80.5 (33.04) |  |  |  |
| Reading Aloud Nonwords (Max=100%) | Baseline | 90.33 (9.29) | 56 (35.67) | 0.07 | 0.50 | 45% |
|  | Outcome | 87.67 (9.29) | 68.25 (40.97) |  |  |  |
| Semantics 2AFC (Max=100%) | Baseline | 93.33 (2.89) | 86.75 (4.27) | 0.79 | 0.015 | 5.6% |
|  | Outcome | 92.67 (4.04) | 87.25 (4.11) |  |  |  |
| Phonology 2AFC (Max=100%) | Baseline | 98.33 (0) | 67.08 (20.48) | 0.04* | 0.60 | 59% |
|  | Outcome | 96.67 (2.89) | 82.50 (15) |  |  |  |
| Orthography 2AFC (Max=100%) | Baseline | 96.67 (0) | 93.33 (4.71) | 0.61 | 0.06 | 7.4% |
|  | Outcome | 97.22 (0.96) | 96.25 (2.85) |  |  |  |
| Manual Dexterity BBT Right Hand | Baseline | 66 (14.24) | 24.25 (17.63) | 0.19 | 0.32 | 24% |
|  | Outcome | 62.67 (7.09) | 29.88 (19.25) |  |  |  |
| Grip Force Right Hand | Baseline | 29.17 (11.06) | 11.85 (8.74) | 0.23 | 0.28 | 20% |
|  | Outcome | 36.87 (6.03) | 12.78 (9.65) |  |  |  |

Note: RCBA – 2AFC – 2 Alternative Forced Choice; BBT – Box and Blocks Test, Reading Comprehension Battery for Aphasia 2<sup>nd</sup> ed., WAB – Western Aphasia. Effect sizes estimated using eta squared ( $\eta^2$ ) should be interpreted as follows: *Small effect*: $\eta^2=0.01$ ; *Medium effect*: $\eta^2=0.06$ ; *Large effect*: $\eta^2=0.14$ ; \* significant at $p<0.05$ , uncorrected.

Across all participants, self-rated study commitment, motivation, and session difficulty were rated significantly higher than 1 for all sessions except for session 2 difficulty (all p’s < 0.005, corrected). In all 3 sessions, commitment and motivation were rated as high and the rating increased from Session 1 to Session 3 (Commitment: Session 1 *M*=6.14, *SD*=1.46; Session 2 *M*=6.57, *SD*=0.77; Session 3 *M*=6.71, *SD*=0.49, Motivation: Session 1 *M*=6.29, *SD*=1.11; Session 2 *M*=6.29, *SD*=0.95; Session 3 *M*=6.57, *SD*=0.53). Difficulty was rated as somewhat easy to neutral (Session 1 *M*=3.71, *SD*=1.98; Session 2 *M*=3, *SD*=1.73; Session 3 *M*=4.29, *SD*=2.06). No significant changes were found in the NFB signal magnitude from before to after the training (See Supplemental Materials).

### Task fMRI

No significant clusters of increased activation were found for stroke participants during the post-compared to the pre-training run. For HCs, significant increases in activation were observed from pre- to post-training during the motor imagery block relative to baseline in the right frontal pole and angular gyrus. Additionally, during word reading relative to baseline, activation increased in the bilateral frontal and the subcallosal cortex. (Table 2).

**Table 2.** Significant Clusters of Increased Task fMRI Activation from Pre- to Post-Training.

| Cluster Index | Cluster Location | Local Maxima (z-value) | MNI Coordinates |  |  |
| --- | --- | --- | --- | --- | --- |
|  |  |  | x | y | z |
| Post-Training > Pre-Training Healthy Controls: NFB Regulation > Baseline |  |  |  |  |  |
| 1 | Right Frontal Pole | 4.19 | 34 | 68 | 0 |
| 2 | Right Angular Gyrus | 3.65 | 64 | -52 | 14 |
| Post-Training > Pre-Training Healthy Controls: Word Reading > Baseline |  |  |  |  |  |
| 1 | Left Superior Frontal and Left Middle Frontal Gyri | 3.03 | -20 | 10 | 50 |
| 2 | Right Middle Frontal Gyrus | 4.35 | 32 | 12 | 50 |
| 3 | Subcallosal Cortex | 5.09 | 0 | 8 | -10 |

When comparing training days 1-3, we found that stroke participants had significantly increased activation on day 2 compared to day 1 for the contrast NFB regulation > baseline in the left basal ganglia, central operculum, and left inferior frontal gyrus (IFG). Similarly, they had greater activation of these areas on day 3 compared to day 1 for NFB regulation > baseline (Table 3). Finally, stroke participants had increased activation on day 3 compared to day 2 in the left inferior and middle temporal, precuneal, and occipital fusiform cortex during nonword reading > baseline. Notably, significant increases in activation for this contrast were observed in the left SMG ROI targeted by the intervention (Fig. 2).

**Table 3.** Significant Clusters of Increased Task fMRI Activation from Day 1 to Day 3 of NFB Training.

| Cluster Index | Cluster Location | Local Maxima (z-value) | MNI Coordinates |  |  |
| --- | --- | --- | --- | --- | --- |
|  |  |  | x | y | z |
| Day 2 > Day 1 Stroke Participants: NFB Regulation > Baseline |  |  |  |  |  |
| 1 | Left Caudate Nucleus | 4.46 | -12 | 10 | 12 |
| 2 | Left Putamen | 3.19 | -32 | -4 | 0 |
| 3 | Left Central Opercular Cortex; Left Inferior Frontal Gyrus, *pars opercularis* | 3.05 | -52 | 4 | -2 |
| 4 | Left Insular Cortex | 3.04 | -34 | -4 | -4 |
| Day 3 > Day 1 Stroke Participants: NFB Regulation > Baseline |  |  |  |  |  |
| 1 | Left Insular Cortex; Frontal Operculum Cortex | 4.46 | -34 | 18 | 4 |
| 2 | Left Inferior Frontal Gyrus, *pars opercularis* | 4.22 | -52 | 10 | 0 |
| Day 3 > Day 2 Stroke Participants: Nonword Reading > Baseline |  |  |  |  |  |
| 1 | Left Inferior Temporal Gyrus, temporooccipital part | 5.47 | -40 | -56 | -2 |
| 2 | Left Precuneus Cortex | 5.01 | -30 | -56 | 8 |
| 3 | Left Occipital Fusiform Gyrus | 4.78 | -26 | -86 | -6 |
| 4 | Left Middle Temporal Gyrus, temporooccipital part | 4.7 | -46 | -54 | -2 |
| Day 3 > Day 2 Healthy Controls: NFB Regulation > Baseline |  |  |  |  |  |
| 1 | Right Frontal Pole | 3.51 | 28 | 42 | -10 |
| 2 | Right Inferior Frontal Gyrus, *pars triangularis* | 3.01 | 42 | 28 | 2 |
| 3 | Right Putamen | 3.00 | 20 | 12 | -4 |
| Day 2 > Day 1 Healthy Controls: Word Reading > Baseline |  |  |  |  |  |
| 1 | Right Nucleus Accumbens; Right Subcallosal Cortex | 4.79 | 10 | 20 | -10 |
| 2 | Left Subcallosal Cortex | 4.27 | -10 | 24 | -8 |
| Day 3 > Day 2 Healthy Controls: Word Reading > Baseline |  |  |  |  |  |
| 1 | Right Supracalcarine Cortex | 3.95 | 12 | -64 | 16 |
| 2 | Left Lingual Gyrus | 3.24 | -8 | -72 | -12 |
| 3 | Right Cuneal Cortex | 3.19 | 6 | -76 | 22 |
| 4 | Left Caudate Nucleus | 3.63 | -20 | 2 | 16 |
| 5 | Left Thalamus | 3.23 | -10 | -10 | 18 |
| 6 | Left Central Opercular Cortex | 3.14 | -38 | -14 | 20 |
| 7 | Right Insular Cortex | 3.82 | 38 | -2 | 2 |
| 8 | Right Precentral Gyrus | 3.4 | 28 | -2 | 20 |
| 9 | Right Thalamus | 3.2 | 16 | -12 | 14 |
| Day 2 > Day 1 Healthy Controls: Nonword Reading > Baseline |  |  |  |  |  |
| 1 | Left Frontal Orbital Cortex | 4.33 | -20 | 20 | -24 |
| 2 | Right Subcallosal Cortex | 3.86 | 8 | 16 | -10 |
| 3 | Right Hippocampus | 4.33 | 30 | -24 | -8 |
| 4 | Right Temporal Occipital Fusiform Cortex | 3.24 | 44 | -40 | -20 |
| 5 | Left Hippocampus | 4.42 | -38 | -42 | 6 |
| *Day 3 > Day 1 Healthy Controls: Nonword Reading > Baseline* |  |  |  |  |  |
| 1 | Right Thalamus | 5.54 | -8 | -18 | 18 |
| 2 | Left Thalamus | 4.61 | -4 | -16 | 18 |

For HCs, activation increased in the right frontal pole, IFG, and putamen on training day 3 compared to day 2 for NFB regulation > baseline. Additionally, on day 2 compared to day 1 participants increased activation in the right nucleus accumbens and bilateral subcallosal cortex for word reading > baseline. For the same contrast, increased activity was found for day 3 relative to day 2 in the right supracalcarine, precentral, cuneal, and insular cortices, bilateral thalamus, left lingual gyrus, central opercular cortex, and caudate nucleus. For nonword reading > baseline, we found increased activity in the left frontal orbital cortex, the right subcallosal and temporal occipital fusiform cortices, and bilateral hippocampus on day 2 compared to day 1 of training. Increased activity in the bilateral thalamus was observed on training day 3 compared to day 1 for the same contrast (Table 3).

### Resting State fMRI

Relative to day 1 of NFB training, rsFC increased bilaterally on day 3 between sensorimotor hand areas (SM hand) and regions of the auditory and cingulo-opercular networks within the same hemisphere. Interhemispheric rsFC also increased, particularly between SM hand areas and between left SM hand and right cingulo-opercular networks. While both HC and stroke participants exhibited this pattern, connectivity strength was weaker in the stroke group (Fig. 3).

**Figure 3.**
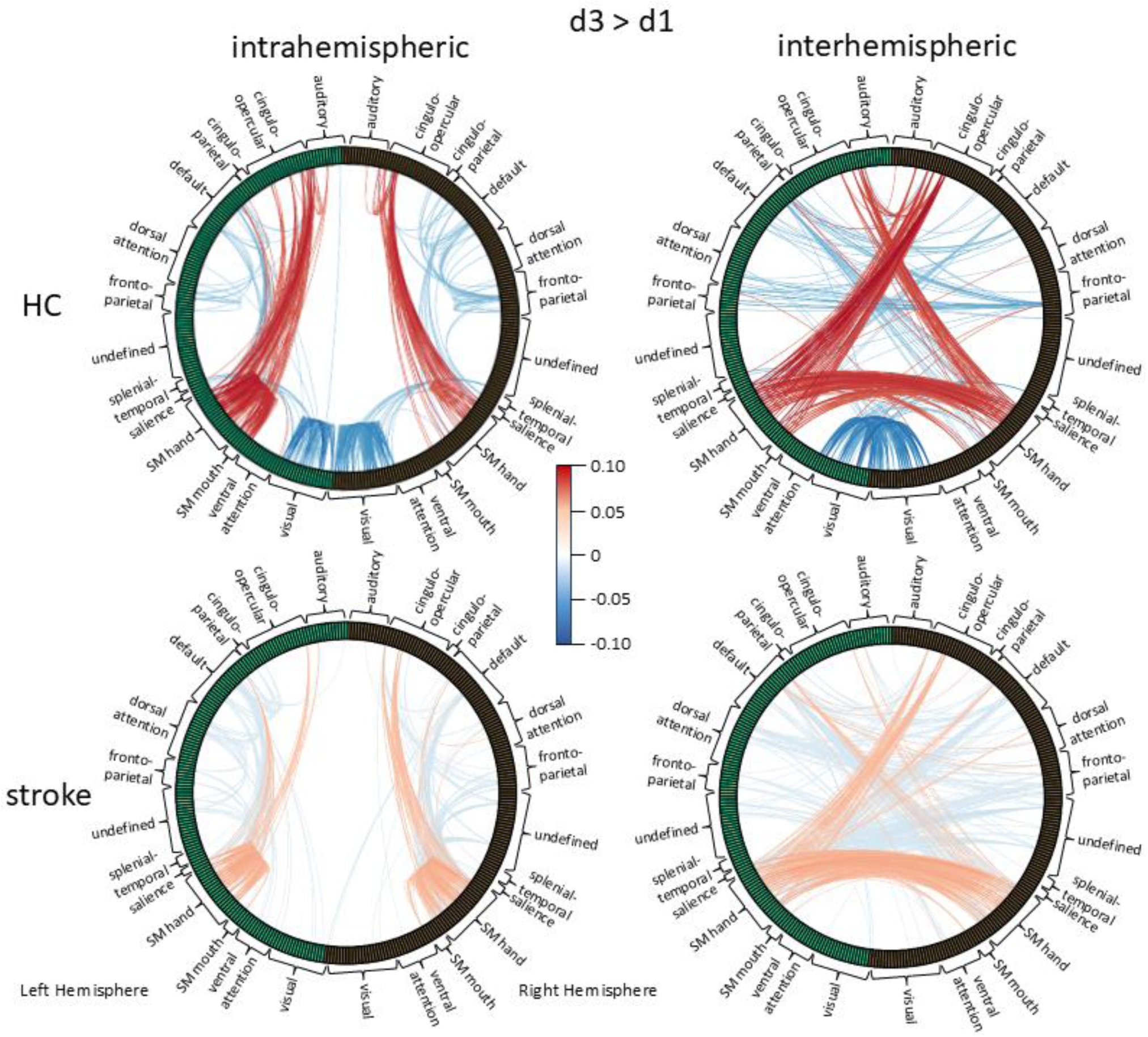
Dynamic resting state functional connectivity across 333 Gordon atlas ROI, comparing brain connectivity following NFB training on day 3 to pretraining resting state scans on day 1. Top 1% of all significant connections are shown. Network labels are from Gordon et al 39 parcellation.

Decreases in rsFC were also observed with some differences between groups. In HC participants, connectivity decreased within the visual cortex (including areas like lateral occipital gyrus, cuneus, lingual and fusiform gyri) and between SM hand, fronto-parietal, cingulo-opercular, and default networks both intra- and inter-hemispherically. Stroke participants showed more widespread decreases in rsFC between fronto-parietal, dorsal attention, default, and cingulo-parietal networks, but without decreases in visual cortex connectivity (Fig. 3).

## 4. Discussion

Reading deficits are common among individuals with aphasia impacting their ability to participate in daily activities^40^. We hypothesized that real-time fMRI NFB could enhance post-stroke reading recovery. Specifically, we targeted the left supramarginal gyrus (SMG), which supports orthography-phonology conversion^19,21^, a process essential for reading and often impaired in aphasia^14,23,24^. Recent studies have shown that among individuals with chronic aphasia reduced CBF in the SMG correlates with diminished language abilities^12^.

In this paper, we present the methodology and preliminary feasibility data for an NFB-based reading intervention evaluating key aspects of the experimental design, including the potential effectiveness of the intervention, participant motivation and commitment, and the appropriateness of the outcome measures.

### 4.1. Behavioral Results

Our preliminary behavioral data indicate that the intervention improved reading and phonological processing in stroke participants, as shown by marked improvements on the RCBA-2 and an experimental phonology test, both with large effect sizes. The RCBA includes sentence and paragraph reading tasks, suggesting generalization beyond single word reading. These results, even within a small sample, highlight specific benefits for stroke participants across a range of reading and phonological tasks. The HC group, which performed at ceiling during baseline and outcome assessment, showed no comparable improvement. However, we cannot rule out the contribution of spontaneous recovery in this subacute stroke sample. Our larger ongoing clinical study will address this limitation by including a non-contingent NFB stroke control group. Furthermore, while we observed numeric group-level increases in NFB signal magnitude from pre- to post-training, these changes were not statistically significant, reflecting individual variability in responsiveness to training, where some participants demonstrated positive increases in NFB signal, while others exhibited no change or even a decrease (Supplemental Fig.2).

The study gathered self-report data on participants’ commitment, motivation, and perceived session difficulty, which provided further insight into the feasibility and acceptability of the intervention. Across all sessions, participants rated their commitment and motivation as high, with both measures increasing slightly from the first to the third session. This trend suggests that participants remained engaged and motivated throughout the intervention. The difficulty of the sessions was rated as somewhat easy to neutral, with a slight increase in perceived difficulty by the third session, indicating that the tasks were challenging yet manageable.

### 4.2. Task fMRI Results

When evaluating brain activity changes after the NFB intervention, we first contrasted pre- and post-training fMRI runs, which were identical to the NFB training runs but without the brain activity feedback. We found no significant increases in brain activity in the stroke participants, suggesting either a lack of power or the absence of sustained brain activity changes beyond the NFB training sessions, warranting further investigation in a larger sample.

In HCs, brain activity increased during NFB regulation and word reading relative to baseline, but these increases occurred outside of the traditional language network^41^. During NFB regulation, activation increased in the right frontal pole, associated with social cognition and emotion regulation^42^, and the right angular gyrus, implicated in episodic memory and egocentric navigation^43^. Word reading activated the left superior and bilateral middle frontal gyri, and the subcallosal cortex, areas implicated in processing unfamiliar text^44^, task switching^45^, goal maintenance^46^ and with individual differences in the ability to control internal thought^47^. Additionally, the subcallosal cortex activation was previously linked with prediction error computation in an audiovisual learning paradigm^48^, memory retrieval of novel scenes^49^, and subsequent memory effects for oddballs relative to control words^50^. These results suggest that NFB training in HCs activates a diverse range of cognitive processes, including executive functions, learning and memory.

We also examined changes in brain activity across training days. In stroke participants, activation increased on day 2 compared to day 1 during NFB regulation relative to baseline in the left basal ganglia, IFG, pars opercularis, central opercular cortex (extending into the auditory cortex), and insula. This activation pattern has been linked with silent reading of motion verbs compared to pseudoverbs^51^, real and imagined stuttering^52^, auditory processing of meaningful sounds^53^, and with processing emotional valence in sounds^54^. The frontal portion of this cluster had also been linked to increased cerebral blood flow related to flexing and extending the fingers of the right hand^55^. The left IFG pars opercularis plays a key role in reading by translating letter representations into their corresponding sounds, in a difficulty sensitive manner. For example, nonwords – being more challenging to process due to their unfamiliarity – activate this area more than real words^19^. Lesions in this region and the left insula have been linked to phonological dyslexia, characterized by difficulty reading nonwords^56^. Comparing day 3 to day 1, we observed similar activation increases in the insula and IFG pars opercularis, indicating that stroke participants engaged areas involved in both language and motor processing throughout training.

In the comparison of day 3 to day 1 we observed increased activation during nonword reading relative to baseline in the left inferior and middle temporal gyri (ITG, MTG), precuneus, and left occipital fusiform cortex. The ITG and MTG are involved in linking conceptual information with word forms, as demonstrated through lesion symptom mapping in aphasia^57,58^. The fusiform cortex, a key part of the reading network, processes written word forms and maps letter-sound and word form-meaning representations^59,60^. While nonwords lack meaning, their processing has been associated with posterior temporal and parietal cortex activation, modulated by task difficulty^61^. Increased precuneus activation may reflect its role in both reading and attention processes, as gray matter volume in this area predicts both skills in children^62^. Thus, as training progressed, stroke participants increased activation of their left reading network including the left SMG—the target region for NFB training. This suggests participants enhanced activation in the SMG and broader reading network, particularly for the more demanding task of nonword reading.

For healthy controls, activation increases for NFB regulation relative to baseline were observed on day 3 compared to day 2 in the right IFG, pars triangularis, frontal pole and putamen, likely reflecting cognitive control mechanisms. Right putamen activation, associated with reward anticipation^63^, may relate to the rewarding effects of positive feedback. Increased activation for word reading relative to baseline included the right nucleus accumbens and bilateral subcallosal cortex on day 2 compared to day 1 and in right supracalcarine, cuneal, and insular cortex, left lingual gyrus, caudate nucleus, and central opercular cortex and bilateral thalamus on day 3 compared to day 2, potentially reflecting reward anticipation^64^, visuo-motor processing^65^, and learning^66^. Increased activity during nonword reading relative to baseline included left frontal orbital, right subcallosal, and occipital fusiform cortex and bilateral hippocampus (day 2 vs day 1) and bilateral thalamus (day 3 vs. day 1). These findings suggest that NFB training engages a broader set of neural circuits in healthy controls, including those involved in reward, cognitive control, and learning.

### 4.3. Resting State fMRI - Dynamic rsFC Results

Following NFB training we observed increased dynamic functional connectivity of the SM hand network^39^, which includes areas associated with motor and sensory processing of hand movements, such as the superior parietal lobule, pre- and postcentral gyri, supplementary motor area (SMA), superior frontal gyrus, and parts of the inferior parietal lobule, including SMG. These regions commonly active during hand motor imagery^67^, became more connected to the cingulo-opercular and auditory networks. The cingulo-opercular network includes motor areas in the SMA, pre-/postcentral gyri, and regions like SMG and IFG, which are associated with reading and language processes. Our findings suggest that practicing hand motor imagery prior to reading during NFB training strengthened functional connectivity supporting hand movements, hearing, and reading. In contrast, we found decreased intra- and interhemispheric connectivity of the fronto-parietal, dorsal attention, default, and SM hand networks, consistent with findings of Zhang and colleagues^68^, who reported reduced default mode network connectivity following imagined motor sequence learning of finger tapping, reflecting decreased introspection and decreased need for cognitive control and integration. We also found decreased visual cortex connectivity in HCs. This is in line with previous studies showing decreased engagement of visual cortex with continued mental imagery practice, which may help minimize interference from external sensory inputs and support the formation of internally generated mental images^69^. The absence of this effect in stroke survivors may reflect their somewhat more challenging experience with motor imagery reflected in post-NFB interviews. Thus, our results are consistent with previous motor imagery literature and indicate that our NFB paradigm modulated the brain networks involved in motor imagery, language, and cognitive control.

## 5. Limitations and Future Directions

To date, only five fMRI NFB stroke rehabilitation studies have been published, including two focused on aphasia with 2-8 stroke participants each. Our study contributes to this emerging field by demonstrating the feasibility of fMRI NFB for post-stroke reading rehabilitation. However, the findings should be interpreted with caution due to limitations such as a male-only sample and the potential influence of spontaneous recovery. Future randomized controlled trials with sham NFB control groups, masking of participants and assessors, and standard-of-care or active control groups will be essential to establish whether fMRI NFB offers additional benefits beyond existing therapies. To address accessibility challenges, future research could explore more cost-effective and portable alternatives to fMRI, such as EEG or fNIRS, for delivering NFB in reading rehabilitation. Despite these limitations, this research marks an important preliminary step toward developing biologically informed interventions for reading deficits in aphasia.

## 8. Data Availability Statement

In accordance with a pre-established data dissemination plan, after project completion, data and the associated documentation will be made available to users upon request under a data-sharing agreement. The agreement mandates: (1) a commitment to using the data only for research purposes and not to identify any individual participant; (2) a commitment to securing the data using appropriate computer technology; and (3) a commitment to destroying or returning the data after analyses are completed.

## 9. Supplemental Material

Supplemental Methods

Supplemental Results

Figures S1-S2

References 69-85

## References

1. Flowers, H. L. et al. Poststroke aphasia frequency, recovery, and outcomes: A systematic review and meta-analysis. Arch. Phys. Med. Rehabil. 97, 2188–2201.e8 (2016).

2. Pedersen, P. M., Vinter, K. & Olsen, T. S. Aphasia after stroke: Type, severity and prognosis: The Copenhagen aphasia study. Cerebrovasc. Dis. 17, 35–43 (2004).

3. Starrfelt, R., Lafsdóttir, R. R. Ó. & Arendt, I. M. Rehabilitation of pure alexia: A review. Neuropsychol. Rehabil. 23, 755–779 (2013).

4. Thibault, R. T., MacPherson, A., Lifshitz, M., Roth, R. R. & Raz, A. Neurofeedback with fMRI: A critical systematic review. Neuroimage 172, 786–807 (2018).

5. Sitaram, R. et al. Closed-loop brain training: The science of neurofeedback. Nat. Rev. Neurosci. 18, 86–100 (2017).

6. Sreedharan, S. et al. Self-regulation of language areas using real-time functional MRI in stroke patients with expressive aphasia. Brain Imaging Behav. 14, 1714–1730 (2020).

7. Sreedharan, S., Arun, K. M., Sylaja, P. N., Kesavadas, C. & Sitaram, R. Functional Connectivity of Language Regions of Stroke Patients with Expressive Aphasia during Real-Time Functional Magnetic Resonance Imaging Based Neurofeedback. Brain Connect. 9, 613–626 (2019).

8. Stockert, A., Kummerer, D. & Saur, D. Insights into early language recovery: from basic principles to practical applications. Aphasiology 39, 517–541 (2016).

9. Thompson, C. K. et al. Intrahemispheric Perfusion in Chronic Stroke-Induced Aphasia. Neural Plast. 236169, 1–15 (2017).

10. Walenski, M. et al. Perilesional Perfusion in Chronic Stroke-Induced Aphasia and Its Response to Behavioral Treatment Interventions. Neurobiol. Lang. 3, 345–363 (2022).

11. Richardson, J. D. et al. Cerebral perfusion in chronic stroke: Implications for lesion-symptom mapping and functional MRI. Behav. Neurol. 24, 117–122 (2011).

12. Ivanova, M. V. et al. Cerebral perfusion in post-stroke aphasia and its relationship to residual language abilities. Brain Commun. 6, 1–17 (2024).

13. Ivanova, M. V. & Pappas, I. Understanding recovery of language after stroke: insights from neurovascular MRI studies. Front. Lang. Sci. 2, (2023).

14. Boukrina, O., Barrett, A. M. M. & Graves, W. W. W. Cerebral perfusion of the left reading network predicts recovery of reading in subacute to chronic stroke. Hum. Brain Mapp. 40, 1–14 (2019).

15. Hillis, A. E. Pharmacological, surgical, and neurovascular interventions to augment acute aphasia recovery. Am. J. Phys. Med. Rehabil. 86, 426–434 (2007).

16. Saur, D. et al. Dynamics of language reorganization after stroke. Brain 129, 1371–1384 (2006).

17. Fridriksson, J. Measuring and inducing brain plasticity in chronic aphasia. J. Commun. Disord. 44, 557–563 (2011).

18. Fridriksson, J., Richardson, J. D., Fillmore, P. & Cai, B. Left hemisphere plasticity and aphasia recovery. Neuroimage 60, 854–863 (2012).

19. Cattinelli, I., Borghese, N. A., Gallucci, M. & Paulesu, E. Reading the reading brain: A new meta-analysis of functional imaging data on reading. J. Neurolinguistics 26, 214–238 (2013).

20. Vogel, A. C., Miezin, F. M., Petersen, S. E. & Schlaggar, B. L. The putative visual word form area is functionally connected to the dorsal attention network. Cereb. Cortex 22, 537–49 (2012).

21. Stoeckel, C., Gough, P. P. M., Watkins, K. K. E. & Devlin, J. J. T. Supramarginal gyrus involvement in visual word recognition. Cortex 45, 1091–1096 (2009).

22. Costanzo, F. et al. Long-lasting improvement following tDCS treatment combined with a training for reading in children and adolescents with dyslexia. Neuropsychologia 130, 38–43 (2019).

23. Boukrina, O., Barrett, A. M., Alexander, E. J., Yao, B. & Graves, W. W. Neurally dissociable cognitive components of reading deficits in subacute stroke. Front. Hum. Neurosci. 9, (2015).

24. Brookshire, C. E., Willson, J. P., Nadeau, S. E., Gonzalez Rothi, L. J. & Kendall, D. L. Frequency, nature, and predictors of alexia in a convenience sample of individuals with chronic aphasia. Aphasiology 1–17 (2014) doi:10.1080/02687038.2014.945389.

25. Andres, M., Pelgrims, B., Olivier, E. & Vannuscorps, G. The left supramarginal gyrus contributes to finger positioning for object use: a neuronavigated transcranial magnetic stimulation study. Eur. J. Neurosci. 46, 2835–2843 (2017).

26. Potok, W., Maskiewicz, A., Króliczak, G. & Marangon, M. The temporal involvement of the left supramarginal gyrus in planning functional grasps: A neuronavigated TMS study. Cortex. 111, 16–34 (2019).

27. Guillot, A. et al. Brain activity during visual versus kinesthetic imagery: An fMRI study. Hum. Brain Mapp. 30, 2157–2172 (2009).

28. Kertesz, A. Western Aphasia Battery Revised. (Pearson, 2007).

29. La Pointe, L. L. & Horner, J. Reading Comprehension Battery for Aphasia. (Pro-Ed, 1998).

30. Sitaram, R. et al. Acquired control of ventral premotor cortex activity by feedback training: An exploratory real-time fMRI and TMS study. Neurorehabil. Neural Repair 26, 256–265 (2012).

31. Robineau, F. et al. Using real-time fMRI neurofeedback to restore right occipital cortex activity in patients with left visuo-spatial neglect: proof-of-principle and preliminary results. Neuropsychol. Rehabil. 29, 339–360 (2019).

32. Oldfield, R. C. The assessment and analysis of handedness: The Edinburgh inventory. Neuropsychologia 9, 97–113 (1971).

33. Kaplan, E., Goodglass, H. & Weintraub, S. Boston Naming Test. (Lea and Febiger, 1983).

34. Mathôt, S., Schreij, D. & Theeuwes, J. OpenSesame: An open-source, graphical experiment builder for the social sciences. Behav. Res. Methods 44, 314–324 (2012).

35. Mathiowetz, V., Volland, G., Kashman, N. & Weber, K. Adult Norms for the Box and Block Test of Manual Dexterity. The American journal of occupational therapy. vol. 39 386–391 (1985).

36. Woolrich, M. W., Ripley, B. D., Brady, M. & Smith, S. M. Temporal autocorrelation in univariate linear modeling of FMRI data. Neuroimage 14, 1370–1386 (2001).

37. Worsley, K. J. Statistical analysis of activation images. in Functional MRI: An Introduction to Methods (ed. Jezzard, P., Matthews, P.M., Smith, S. M.) (Oxford University Press, 2001).

38. Pruim, R. H. R. et al. ICA-AROMA: A robust ICA-based strategy for removing motion artifacts from fMRI data. Neuroimage 112, 267–277 (2015).

39. Gordon, E. M. et al. Generation and Evaluation of a Cortical Area Parcellation from Resting-State Correlations. Cereb. Cortex 26, 288–303 (2016).

40. Knollman-Porter, K., Wallace, S. E., Hux, K., Brown, J. & Long, C. Reading experiences and use of supports by people with chronic aphasia. Aphasiology 29, 1448–1472 (2015).

41. Fedorenko, E., Hsieh, P. J., Nieto-Castañón, A., Whitfield-Gabrieli, S. & Kanwisher, N. New method for fMRI investigations of language: Defining ROIs functionally in individual subjects. J. Neurophysiol. 104, 1177–1194 (2010).

42. Ray, K. L. et al. Co-activation based parcellation of the human frontal pole. Neuroimage 123, 200–211 (2015).

43. Teghil, A., Bonavita, A., Guariglia, C. & Boccia, M. Commonalities and specificities between environmental navigation and autobiographical memory: A synthesis and a theoretical perspective. Neurosci. Biobehav. Rev. 127, 928–945 (2021).

44. Buchweitz, A., Mason, R. A., Meschyan, G., Keller, T. A. & Just, M. A. Modulation of cortical activity during comprehension of familiar and unfamiliar text topics in speed reading and speed listening. Brain Lang. 139, 49–57 (2014).

45. Rodríguez-Pujadas, A. et al. Bilinguals Use Language-Control Brain Areas More Than Monolinguals to Perform Non-Linguistic Switching Tasks. PLoS One 8, (2013).

46. Spiers, H. J. & Maguire, E. A. Thoughts, behaviour, and brain dynamics during navigation in the real world. Neuroimage 31, 1826–1840 (2006).

47. Banich, M. T., Mackiewicz, K. L., Depue, B. E. & Burgess, G. C. Multiple modes of clearing one’s mind of current thoughts: Overlapping and distinct neural systems. Neuropsychologia 69, 105–117 (2015).

48. Iglesias, S., et al. Editorial Note to: Hierarchical Prediction Errors in Midbrain and Basal Forebrain during Sensory Learning. Neuron 101, 1195 (2019).

49. Rausch, V. H., Bauch, E. M. & Bunzeck, N. White noise imporves learning by modulating activity in dopaminergic midbrain regions and right superior temporal sulcus. J. Cogn. Neurosci. 26, 1469–1480 (2014).

50. Strange, B. A. et al. Dopamine receptor 4 promoter polymorphism modulates memory and neuronal responses to salience. Neuroimage 922–931 (2014).

51. Rodríguez-Ferreiro, J., Gennari, S. P., Davies, R. & Cuetos, F. Neural correlates of abstract verb processing. J. Cogn. Neurosci. 23, 106–118 (2011).

52. Wymbs, N. F., Ingham, R. J., Ingham, J. C., Paolini, K. E. & Grafton, S. T. Individual differences in neural regions functionally related to real and imagined stuttering. Brain Lang. 124, 153–164 (2013).

53. Lima, C. F. et al. Feel the noise: Relating individual differences in auditory imagery to the structure and function of sensorimotor systems. Cereb. Cortex 25, 4638–4650 (2015).

54. Viinikainen, M., Kätsyri, J. & Sams, M. Representation of perceived sound valence in the human brain. Hum. Brain Mapp. 33, 2295–2305 (2012).

55. Fink, G. R., Frackowiak, R. S. J., Pietrzyk, U. & Passingham, R. E. Multiple nonprimary motor areas in the human cortex. J. Neurophysiol. 77, 2164–2174 (1997).

56. Ripamonti, E. et al. The anatomical foundations of acquired reading disorders: A neuropsychological verification of the dual-route model of reading. Brain Lang. 134, 44–67 (2014).

57. Acres, K., Taylor, K. I., Moss, H. E., Stamatakis, E. a & Tyler, L. K. Complementary hemispheric asymmetries in object naming and recognition: a voxel-based correlational study. Neuropsychologia 47, 1836–43 (2009).

58. Henseler, I., Regenbrecht, F. & Obrig, H. Lesion correlates of patholinguistic profiles in chronic aphasia: comparisons of syndrome-, modality- and symptom-level assessment. Brain 137, 918–30 (2014).

59. Seghier, M. L., Lee, H. L., Schofield, T., Ellis, C. L. & Price, C. J. Inter-subject variability in the use of two different neuronal networks for reading aloud familiar words. Neuroimage 42, 1226–36 (2008).

60. Graves, W. W., Desai, R., Humphries, C., Seidenberg, M. S. & Binder, J. R. Neural systems for reading aloud: a multiparametric approach. Cereb. Cortex 20, 1799–815 (2010).

61. Graves, W. W., Boukrina, O., Mattheiss, S. R., Alexander, E. J. & Baillet, S. Reversing the standard neural signature of the word-nonword distinction. J. Cogn. Neurosci. 29, (2017).

62. Lee, M. M., Drury, B. C., McGrath, L. M. & Stoodley, C. J. Shared grey matter correlates of reading and attention. Brain Lang. 237, (2023).

63. Kirk, U., Brown, K. W. & Downar, J. Adaptive neural reward processing during anticipation and receipt of monetary rewards in mindfulness meditators. Soc. Cogn. Affect. Neurosci. 10, 752–759 (2015).

64. Knutson, B., Adams, C. M., Fong, G. W. & Hommer, D. Anticipation of increasing monetary reward selectively recruits nucleus accumbens. J. Neurosci. 21, 1–5 (2001).

65. Kim, E. et al. Deciphering Functional Connectivity Differences Between Motor Imagery and Execution of Target-Oriented Grasping. Brain Topogr. 36, 433–446 (2023).

66. Tricomi, E., Delgado, M. R., McCandliss, B. D., McClelland, J. L. & Fiez, J. a. Performance feedback drives caudate activation in a phonological learning task. J. Cogn. Neurosci. 18, 1029–43 (2006).

67. Saiote, C. et al. Resting-state functional connectivity and motor imagery brain activation. Hum. Brain Mapp. 37, 3847–3857 (2016).

68. Zhang, H. et al. Motor imagery learning modulates functional connectivity of multiple brain systems in resting state. PLoS One 9, (2014).

69. Koush, Y., Masala, N., Scharnowski, F. & Van De Ville, D. Data-driven tensor independent component analysis for model-based connectivity neurofeedback. Neuroimage 184, 214–226 (2019).

